# Automatic analysis system of COVID-19 radiographic lung images (XrayCoviDetector)

**DOI:** 10.1101/2020.08.20.20178723

**Authors:** Nicolas Schlotterbeck, Carlos E. Montoya, Patricia Bitar, Jorge A. Fuentes, Victor Dinamarca, Gonzalo M. Rojas, Marcelo Galvez

## Abstract

COVID-19 is a pandemic infectious disease caused by the SARS-CoV-2 virus, having reached more than 210 countries and territories. It produces symptoms such as fever, dry cough, dyspnea, fatigue, pneumonia, and radiological manifestations.

The most common reported RX and CT findings include lung consolidation and ground-glass opacities.

In this paper, we describe a machine learning-based system (XrayCoviDetector; until the image has a size www.covidetector.net), that detects automatically, the probability that a thorax radiological image includes COVID-19 lung patterns.

XrayCoviDetector has an accuracy of 0.93, a sensitivity of 0.96, and a specificity of 0.90.

## 1. Introduction

COVID-19 (acronym of coronavirus disease 2019), also known as coronavirus disease or coronavirus, is an infectious disease caused by the SARS-CoV-2 virus (Wu, 2020; Huang, 2020). It was first detected in the city of Wuhan, China in December 2019 (Wu, 2020; Huang, 2020). Having reached more than 210 countries, areas, and territories (WHO, 2020), the World Health Organization declared it a pandemic on March 11, 2020 (WHO, 2020).

It produces flu-like symptoms including fever, dry cough, dyspnea, myalgia, and fatigue. In severe cases, it is characterized by pneumonia, acute respiratory distress syndrome, sepsis, and septic shock (Huang, 2020).

COVID-19 has radiological manifestations, even in asymptomatic patients, and in certain cases before a positive real-time reverse transcription-polymerase chain reaction (RT-PCR) test. Radiological features have already been used to identify high-risk patients early, improving the prognosis (and reducing the need for invasive mechanical ventilation) by being able to establish monitoring and early intensive management.

## 2. Material and Methods

3805 radiography of normal, pneumonia not COVID-19, and COVID-19 type pneumonia was used. The database is made up of three sources:

To minimize false-positive errors, the images of three databases (Table 1) were reviewed, validated, and selected by expert radiologists (obtaining the number of images detailed in Table 1), since not all patients show distinguishable patterns on their chest radiographs. It should be noted that the first set (item 1 in Table 1) is RSNA images and is validated for a pre-pandemic Kaggle competition (patients without COVID).

**Table 1:** Image databases used to train and validate the AI system.

| # | ORIGIN | TYPE | QUANTITY |
| --- | --- | --- | --- |
| 1 | RSNA Pneumonia Detection Challenge (https://www.kaggle.com/c/rsna-pneumonia-detection-challenge) | Normal, and non-COVID-19 pneumonia | 3014 |
| 2 | Clínica las Condes | COVID-19+ | 573 |
| 3 | COVID-19 image data collection (https://github.com/ieee8023/covid-chestxray-dataset) (Maguolo, 2020) | COVID-19+ | 218 |

Being a small data set, two sets were randomly formed. One set was used to train the model and the other to validate it (not test set was formed). See Table 2.

**Table 2:**
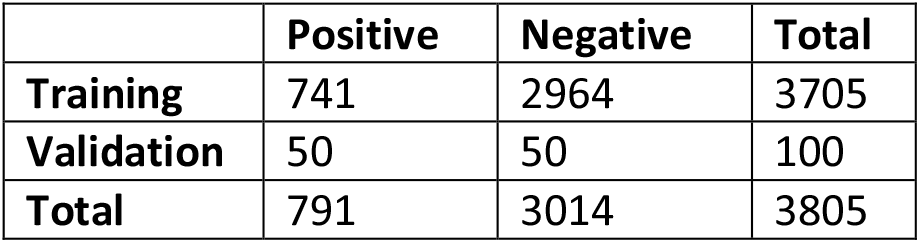
Quantity of RX images used to train and validate the network model. Positive: COVID-19 images, Negative: normal and non-COVID-19 pneumonia images.

|  | Positive | Negative | Total |
| --- | --- | --- | --- |
| **Training** | 741 | 2964 | 3705 |
| **Validation** | 50 | 50 | 100 |
| **Total** | 791 | 3014 | 3805 |

In the training set, we have four times fewer cases of COVID, so different weights are introduced to each category to measure the error in the training of the model (in addition to passing the data in large batches of 100 images each, such that contain multiple COVID images at each training step).

To measure the precision of the model (correct results over totals), note that the validation set is balanced so that there are equal numbers of positive and negative cases (Table 2).

### Preprocessing

#### U-Net

To avoid bias, by date or origin of images, annotations are removed from images using an automatic segmentation system. A U-Net-type AI architecture (Ronneberger et al., 2015) was trained to perform automatic segmentation of only the lung before performing the classification (Figure 1, 2, 3). The data was grayscale normalized before the segmentation.

**Figure 1:**
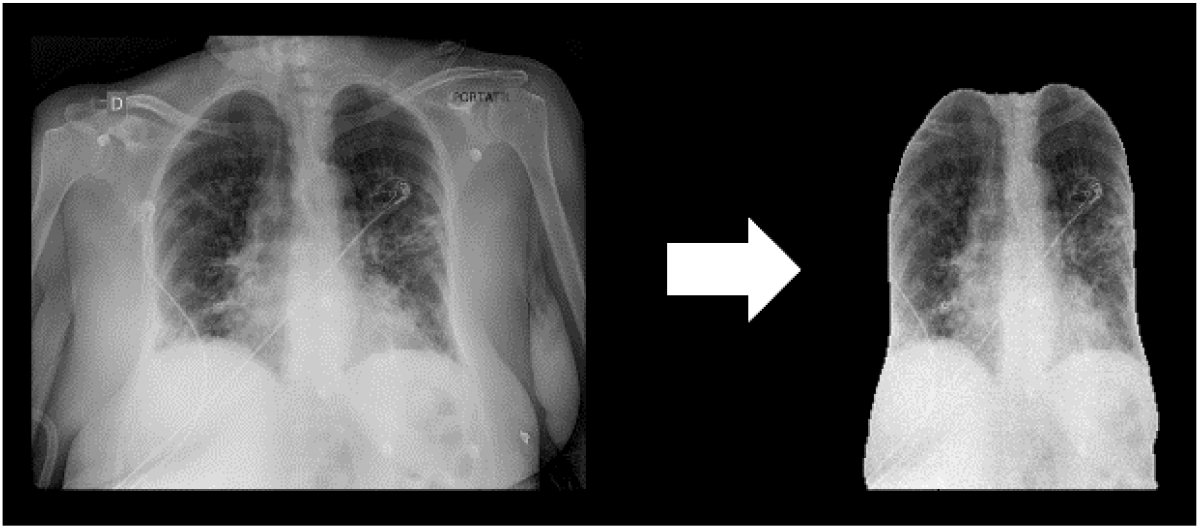
UNet-type AI architecture was used to automatically segment the lung. (**A**) RX original lung image, (**B**) segmented lung.

**Figure 2:**
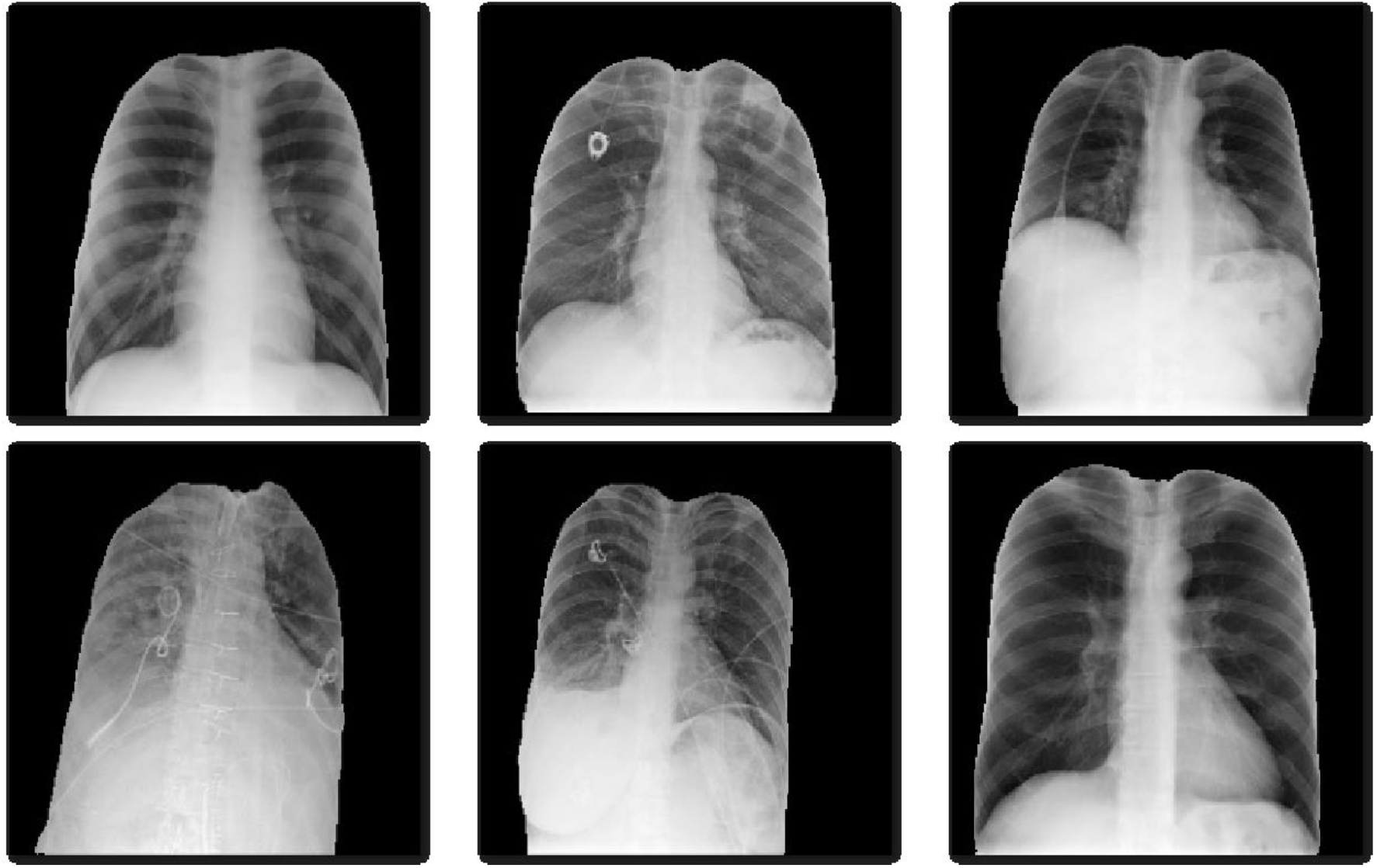
Different RX images of segmented lungs using the U-Net-type AI model.

**Figure 3:**
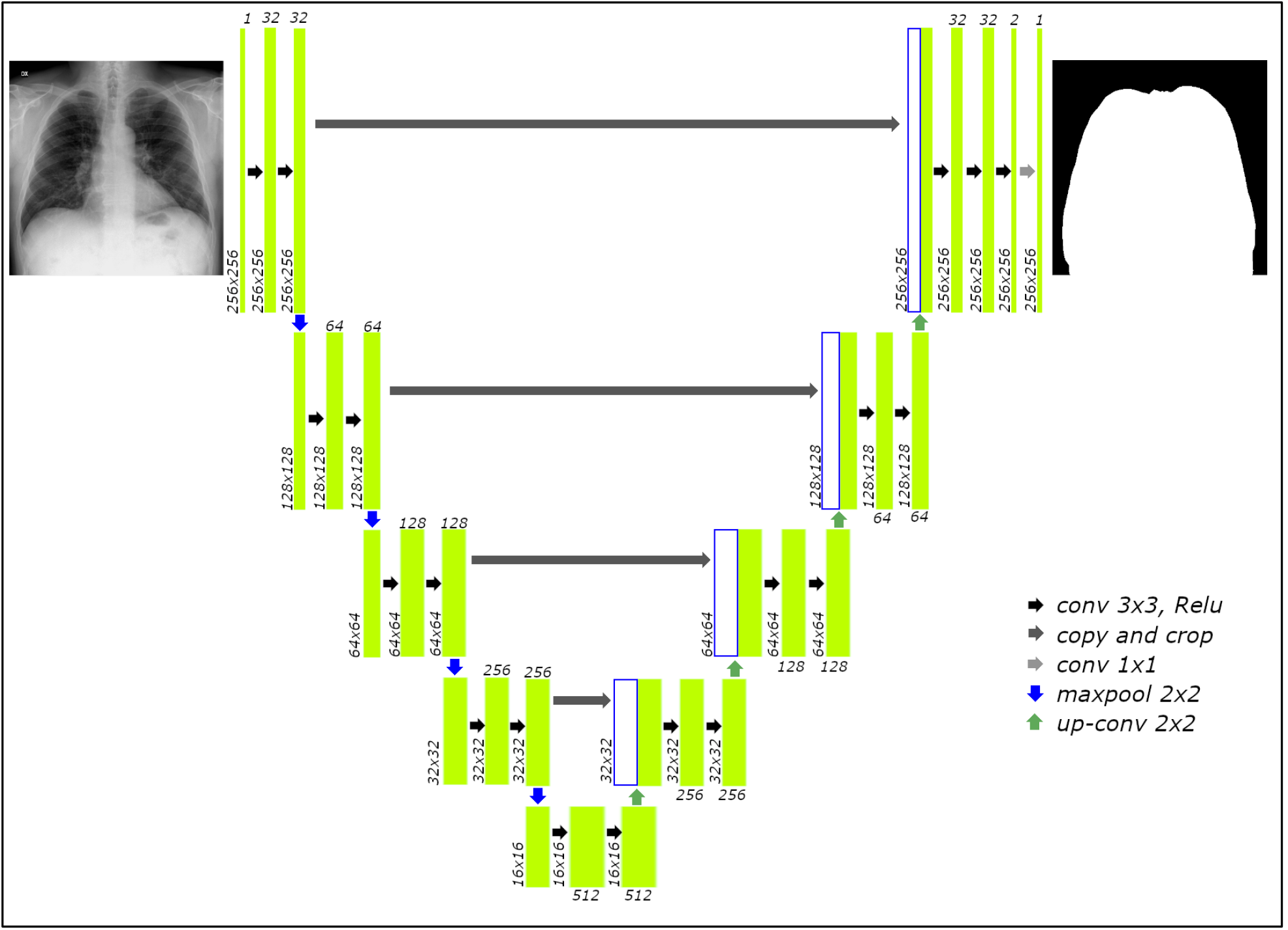
U-net architecture used to segment the lung from DX images.

U-net architecture is shown in Figure 3. It consists of contraction (encoder network) and expansion path(decoder network).

#### Contraction path

The 256×256×1 input image is passed through two 3×3 convolutional layers. Then, the image is downsampled using 2×2 max-pooling layers. This process is repeated until the image has a size of 16×16×512.

#### Expansion path

Then instead of downsampling, the image is sent through a 2×2 deconvolutional layer (up-Conv 2×2; Fig 3) and concatenated consecutively with a cropped version of the previous feature map, and similarly, the feature map is sent through 3×3 convolutional layers. The process is repeated until an image of size 256×256×2 (Fig ×) was got and a 1×1 convolution layer was applied to get a 256×256×1 sized output (1 class: lung).

#### Data augmentation

The data was expanded by applying different transformations randomly. In Table 3, the transformations for image segmentation model, and in Table 4 the transformations for classification model are shown.

**Table 3:** Keras Augmentation parameters for transformations that will be applied to each radiological image for segmentation model (Fig. 3).

| TRANSFORMATION | RANGE |
| --- | --- |
| Shear | 0.05 |
| Rotation | 0.2 |
| Zoom | 0.05 |
| Width shift | 0.05 |
| Height shift | 0.05 |
| Horizontal flip | True |
| fill_mode | nearest |

**Table 3:** Transformations that will be applied to each radiological image to augment the data set for classification model (Fig. 5).

| TRANSFORMATION | RANGE |
| --- | --- |
| Shear | 0.05 |
| Rotation | 0.2 |
| Zoom | 0.2 |
| Width shift | 0.2 |
| Height shift | 0.2 |
| Brightness | [0.1, 1.9] |
| Horizontal flip | True |

**Table 4:**
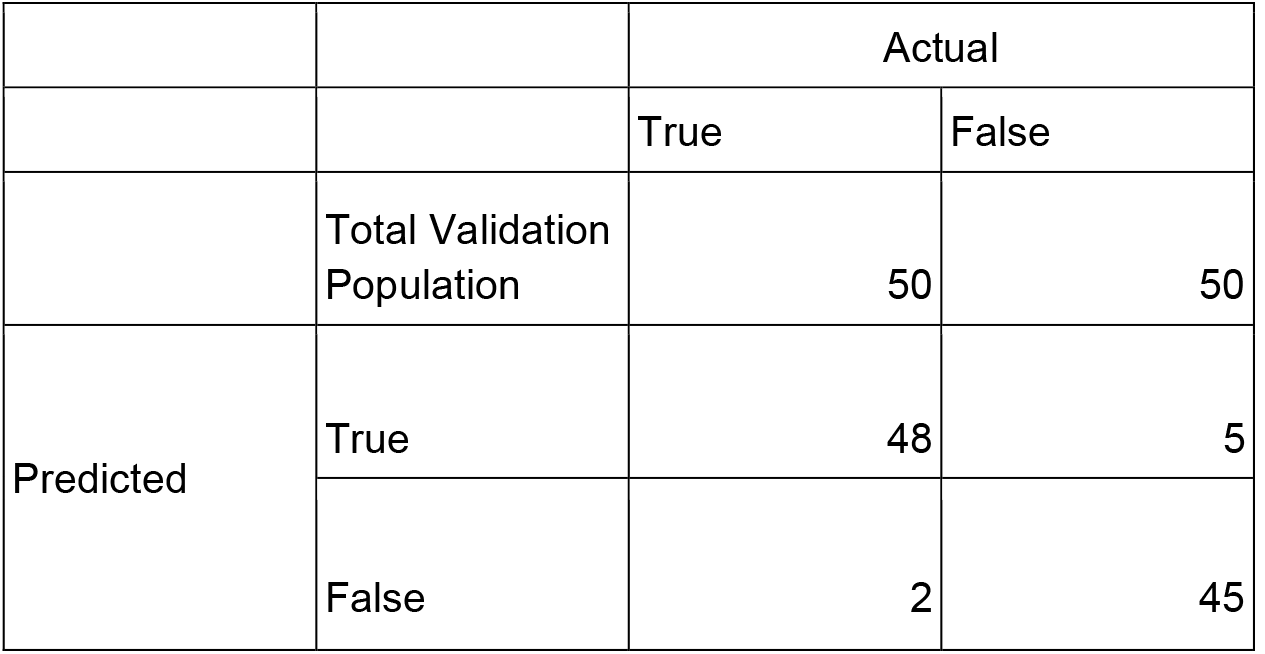
Confusion matrix for the validation set.

The transformations (Table 3) produce greater variability to the data and help the model to improve generalization (so that it responds better with new data and with different distributions, trying to make it more robust and reliable). See Figure 4.

**Figure 4:**
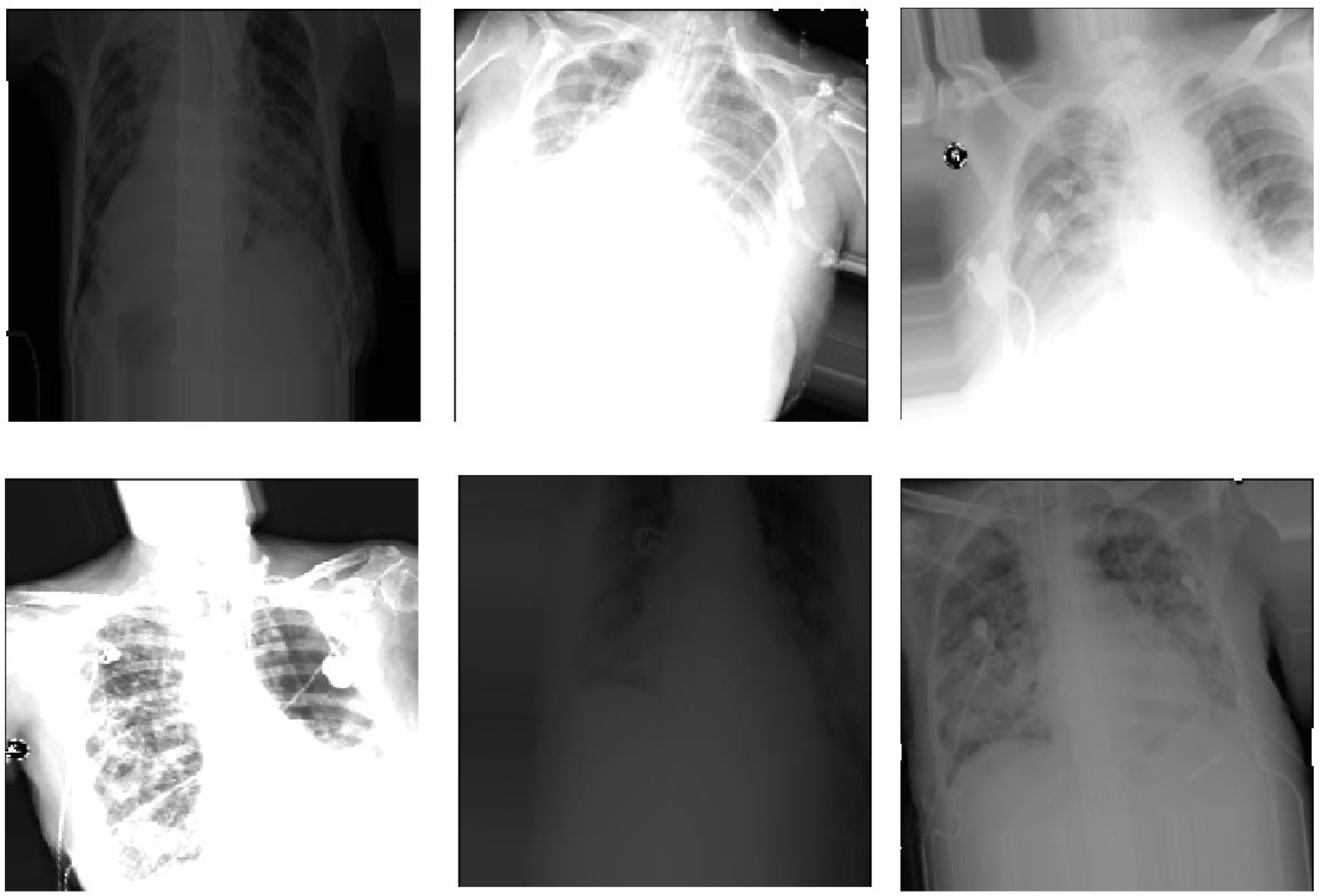
Example of data augmented by applying different transformations (Table 3).

#### Classification model

Given that the data sets used are relatively small, it was decided to use transfer learning from a pre-trained classic VGG-16 model (a convolutional neural network that is 16 layers deep; Fig 5) in the first convolutional layers with fixed weights obtained on ImageNet images (http://www.image-net.org/). ImageNet set contains 120 categories of images of all kinds, not from the medical area, so in the first fixed convolutional layers, the model contains the filters for the relevant characteristics typical of any image (for example, edge detection).

**Figure 5:**
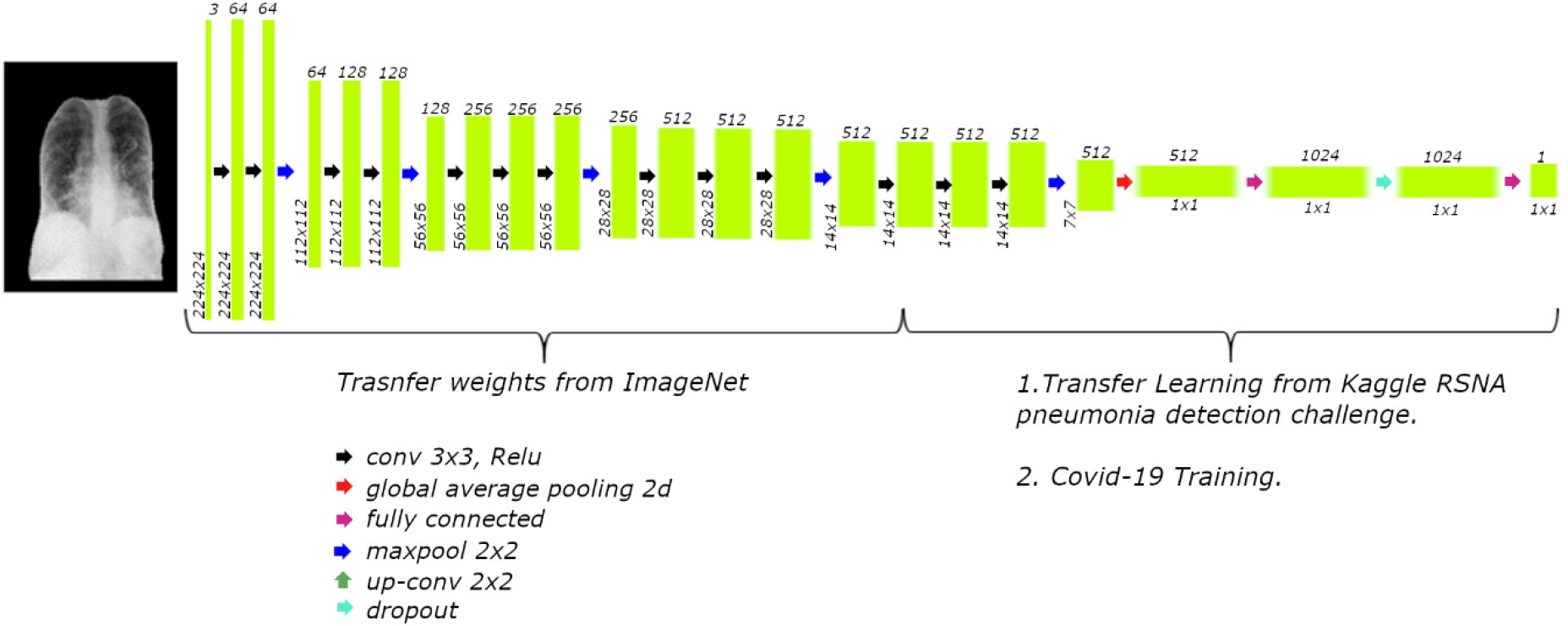
GG16 architecture used to predict COVID19 using chest radiography previously segmented.

For the unfrozen layers of the model, it is necessary to change the last dense layers to be of two categories. The unfreeze layers are trained for a slightly different problem. This time a problem from the medical area is used, with chest radiographs for the detection of pneumonia with over twelve thousand images from a Kaggle competition (with images very similar to the final problem to be solved, but where there are more images. RSNA Pneumonia Detection Challenge; https://www.kaggle.com/c/rsna-pneumonia-detection-challenge). Also, since COVID can cause a type of pneumonia, the problems are similar and help the development of the system.

Finally, you train on the final problem. During training, some neurons are randomly removed (given the lack of data, to avoid overfitting the training set and poor generalization).

#### Implementation

Using Amazon Web Services (AWS; https://aws.amazon.com) XrayCoviDetector was implemented. A website, and the complete neural network. Using Amazon EC2 (https://aws.amazon.com/es/ec2/), Amazon S3 (https://aws.amazon.com/es/s3/) for the storage and web site, Amazon SageMaker (https://aws.amazon.com/es/sagemaker/) and Amazon TensorFlow (https://aws.amazon.com/es/tensorflow/) to create and implement the neural networks used in this project.

## 3. Results

A website was created: http://www.covidetector.net (XrayCoviDetector; Fig 6).

**Figure 6:**
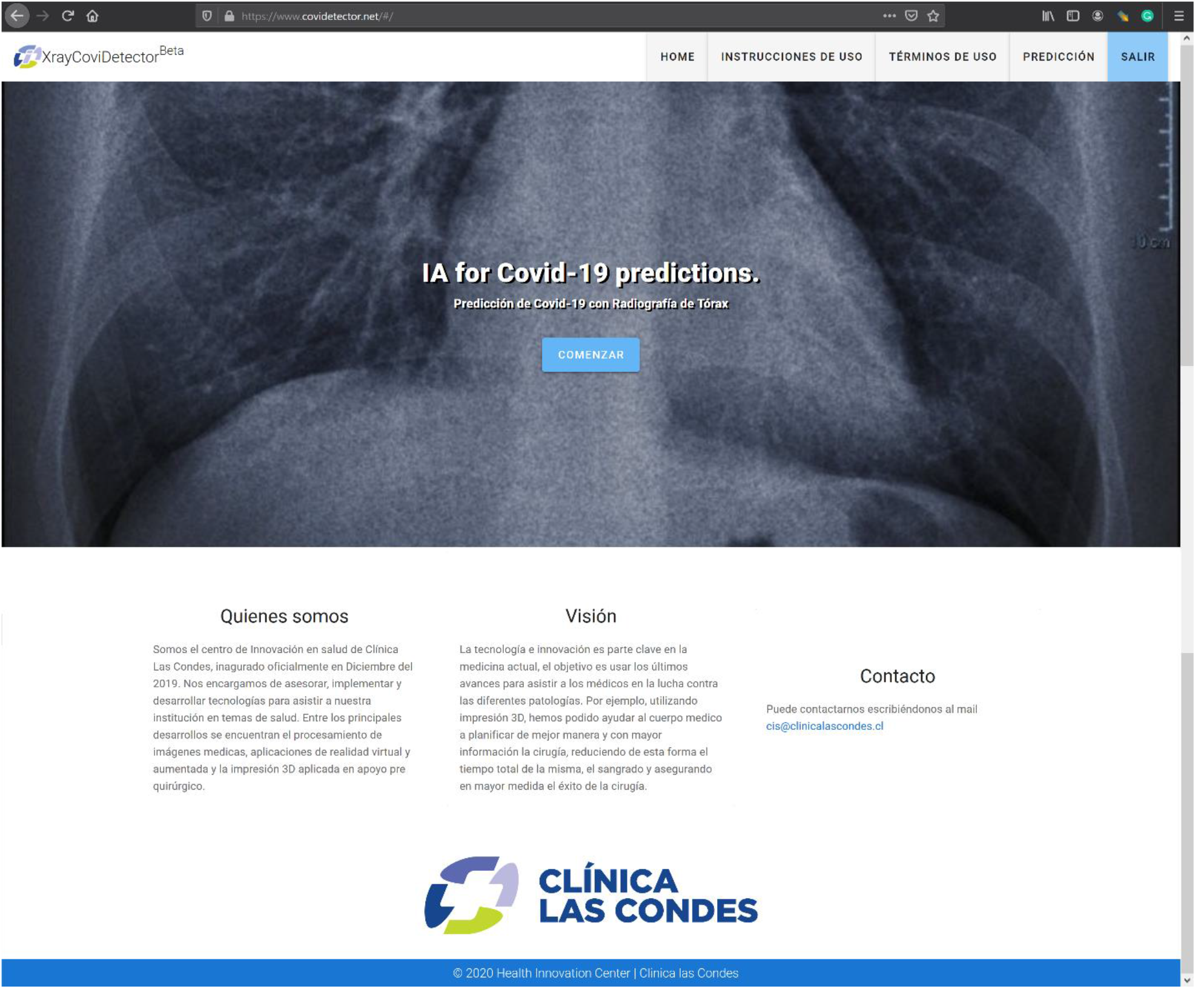
Home page of XrayCoviDetector.

To login to XrayCoviDetector, the user must input his email and password (Fig 7A), and register a new user the complete name, email, password, and must click the checkbox to know and accept the conditions of use (Fig 7B).

**Figure 7:**
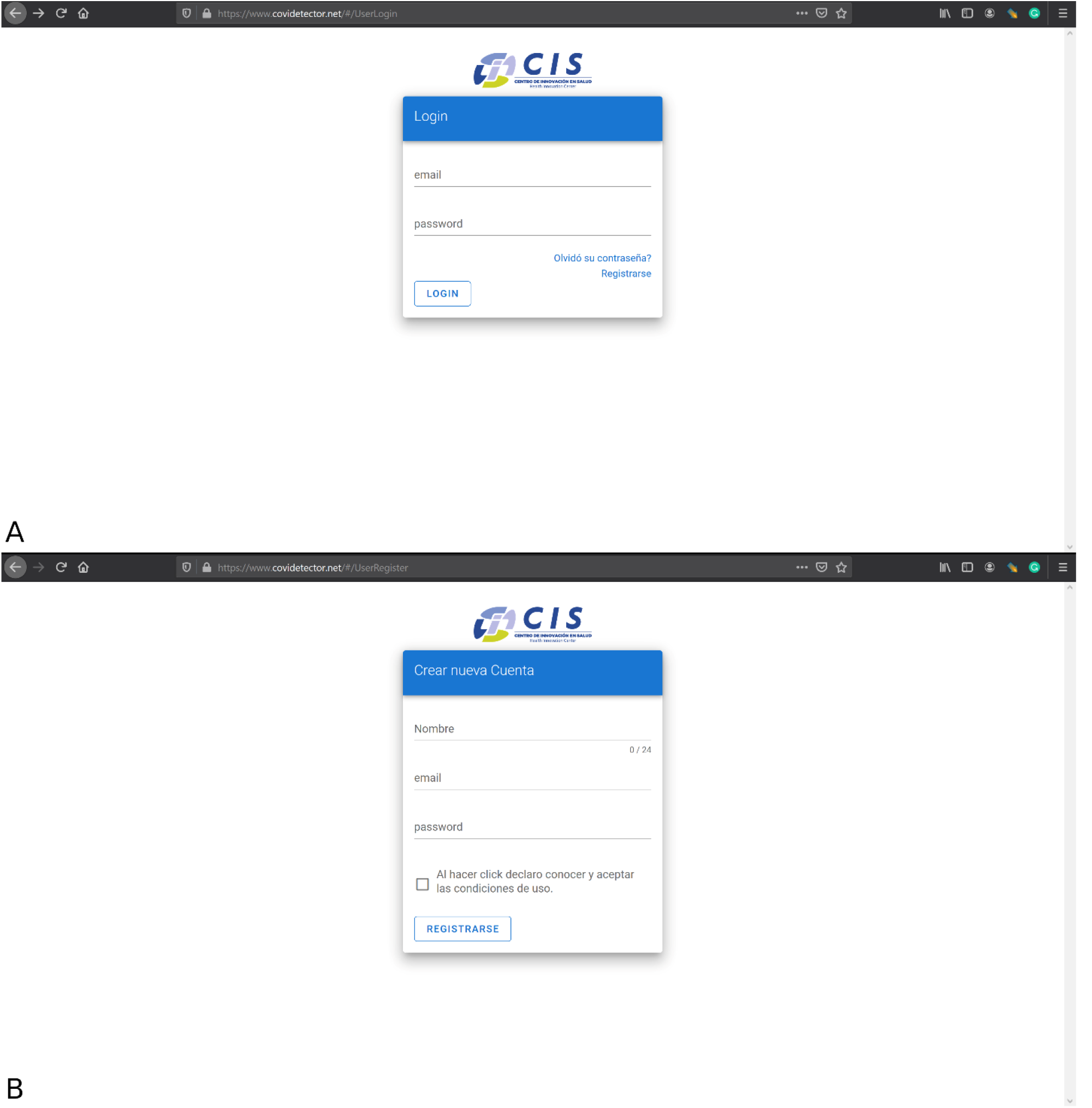
(A) Login web page, and (B) User registration webpage.

To upload a case, the age, sex, and a PNG or JPG chest image of the patient must be selected (Figure 8). Then press the “ENVIAR” button.

**Figure 8:**
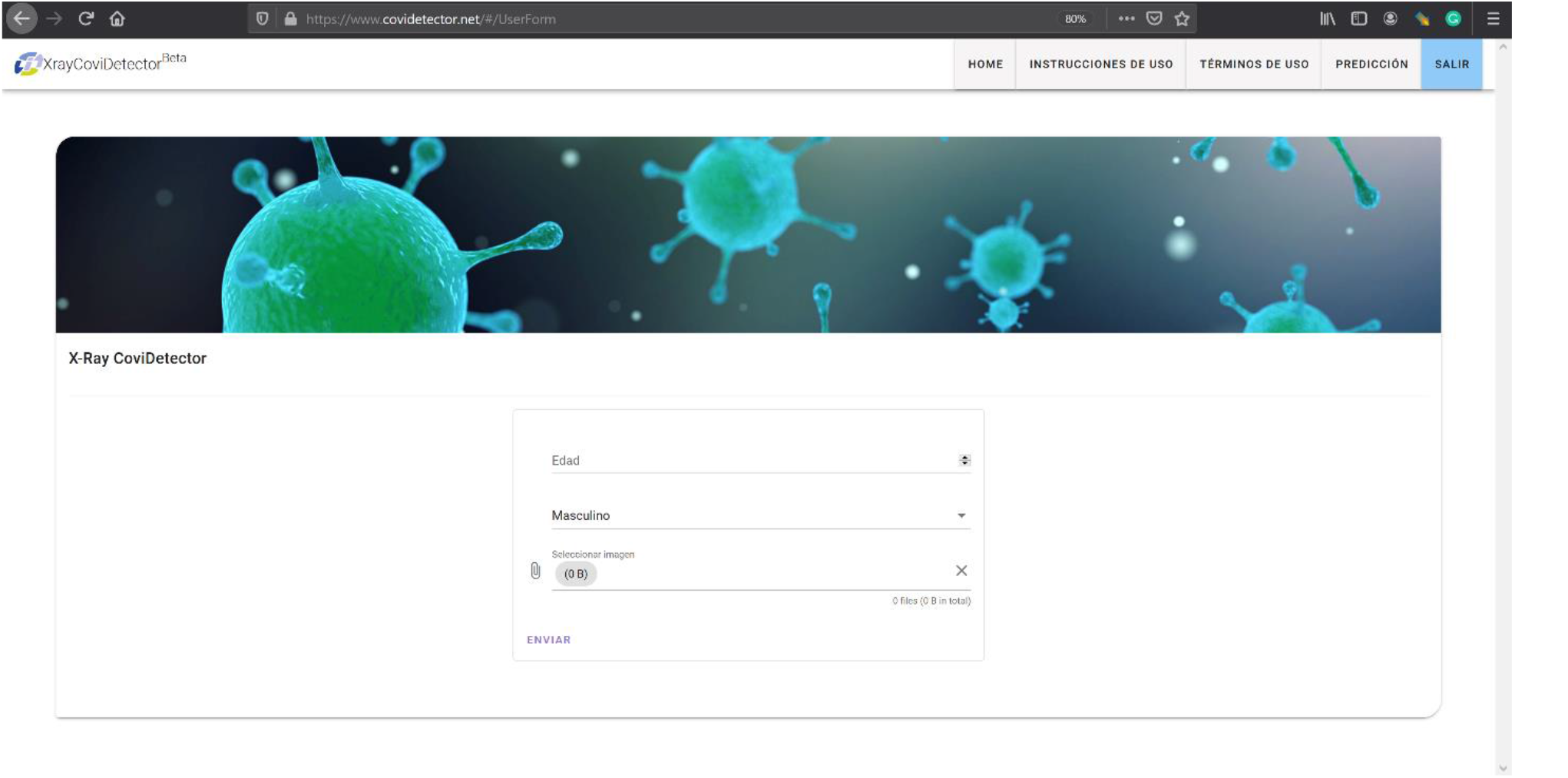
Web page to upload a case. Age, sex, and PNG or JPG chest image files must be selected to upload by pressing the “ENVIAR” button.

Later, the user will receive an email from with the result of the analysis of RX images of the patient. Two possible sentences appear in the received email: “se detectan posibles patrones de COVID-19” (“possible COVID-19 patterns were detected”) or “no se detectaron patrones de COVID-19” (“COVID-19 patterns not detected”). See Figure 9 and Figure 10.

**Figure 9:**
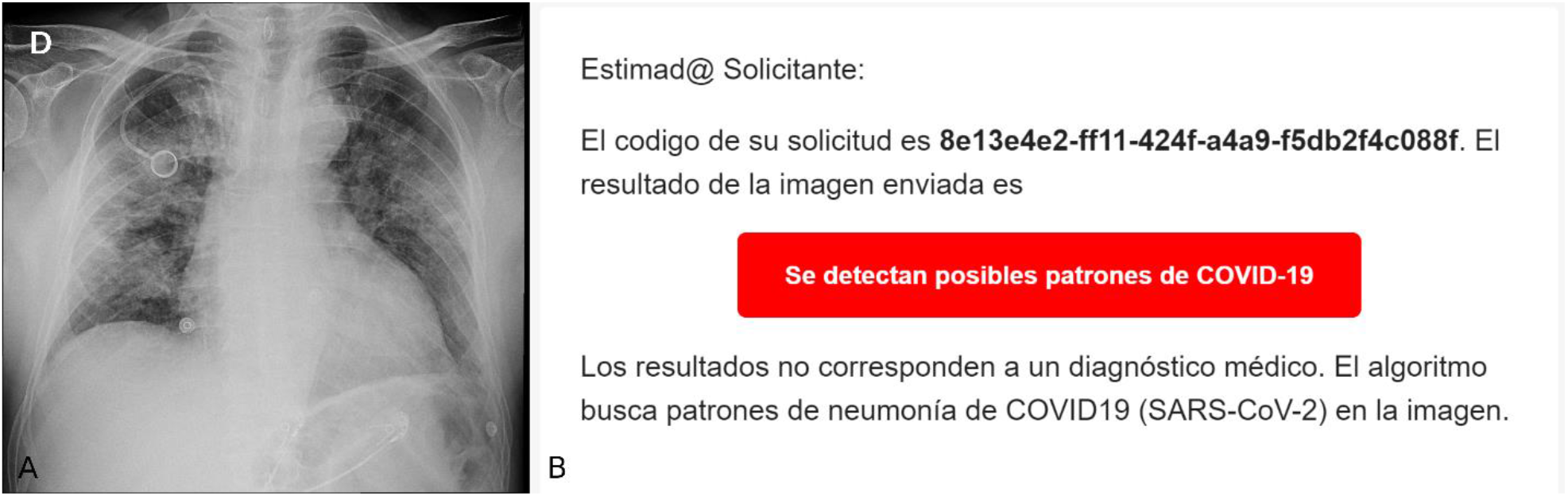
(A) COVID-19 DX image of male 75 years old patient (Parenchymal opacities with alveolar appearance and bilateral ground-glass opacity, mainly in the middle third, to a greater extent to the right), (B) e-mail received with the analysis of the COVID-19 case (sentence “possible COVID-19 patterns was detected”).

**Figure 10:**
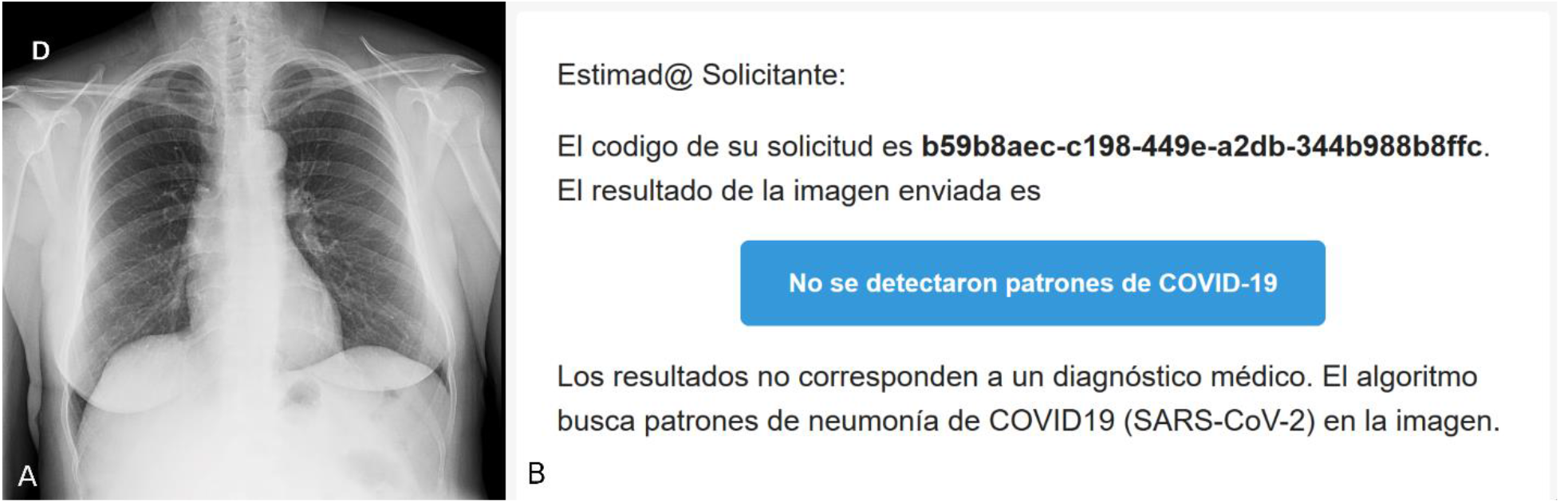
(A) Lung DX image of a healthy 51 years old female patient, (B) e-mail received with the analysis of healthy patient (“COVID-19 patterns not detected”).

The results of the validation set, even distorting the images, are shown in a confusion matrix (Table 4).

### Accuracy

Refers to how close to the actual value the measured value is. High accuracy means that there is a small difference between the predicted result of XrayCoviDetector and the actual positive (RX lung with COVID-19 pneumonia) (Griffiths, 2009);

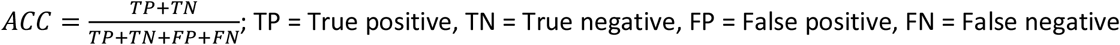

### Sensitivity

(True Positive Rate): measures the proportion of actual positives (RX lung with COVID-19 pneumonia) that are correctly identified as such by XrayCoviDetector (Griffiths, 2009):

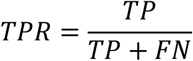

### Specificity

(True Negative Rate): measures the proportion of actual negatives (RX lung without COVID-19 pneumonia) that are correctly identified as such by XrayCoviDetector (Griffiths, 2009):

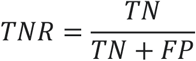

Using data of Table 4, the accuracy is equal to 0.93, sensitivity = 0.96, and specificity = 0.90.

To verify that the system does not have any kind of bias related to the origin of the data, the system was tested using 66 CLC origin only images, obtaining an accuracy of 0.92.

## 4. Discussion

In this paper, a COVID-19 lung pattern automatic detection system using RX images was described. XrayCoviDetector is a worldwide accessible web system from a computer or a mobile device, easy to use that send the results through email in a couple of seconds.

To train and validate the system, images from different origin databases, and probably different technical characteristics have been used (Table 1). With these data, a high accuracy (0.93) was obtained. To assess whether the accuracy is maintained, XrayCoviDetector was measured using CLC acquired images, obtaining an accuracy of 0.92. Being able to conclude that the trained neural network is robust, and the results will not depend significantly on the characteristics of the images or the X-ray equipment used.

XrayCoviDetector is a fast and fully automatic chest X-Ray analysis web system that detects COVID-19 pneumonia patterns. Pieces of X-Ray equipment are common in medical centers and hospitals worldwide, but PCR (the gold-standard COVID-19 diagnosis exam) is not as common. Therefore, X-Ray can be considered as a complementary support exam, and XrayCoviDetector supports the non-existence of a COVID-19 expert radiologist in the medical center.

XrayCoviDetector may be less effective in detecting lung disease patterns in early stages than in more advanced stages. This is because there are fewer images of the early stages in the training set than in more advanced stages.

It is important to note that many of the COVID-19 images used for this system correspond to the same patient on different days (so some of the images look similar but highlighting that they do not they are identical). To reduce this problem, they are randomly sorted and passed in separate groups during training (in addition to random distortion on augmentation).

## Data Availability

None

## Notes

### Competing Interest Statement

The authors have declared no competing interest.

### Author Declarations

Approved by the Research Ethics Committee of Clinica las Condes, Santiago, Chile.

## References

Chung, M., Bernheim, A., Mei, X., Zhang, N., Huang, M., Zeng, X., Cui, J., Xu, W., Yang, Y., Fayad, Z. A., Jacobi, A., Li, K., Li, S., & Shan, H. (2020). CT Imaging Features of 2019 Novel Coronavirus (2019-nCoV). Radiology, 295(1), 202–207. https://doi.org/10.1148/radiol.2020200230

Griffiths, D. (2009). Head First Statistics: A Brain-Friendly Guide. O’Reilly Medica Inc., CA, USA. ISBN: 978-0-596-52758-7.

Huang, C., Wang, Y., Li, X., Ren, L., Zhao, J., Hu, Y., Zhang, L., Fan, G., Xu, J., Gu, X., Cheng, Z., Yu, T., Xia, J., Wei, Y., Wu, W., Xie, X., Yin, W., Li, H., Liu, M., Xiao, Y., Gao, H., Guo, L., Xie, J., Wang, G., Jiang, R., Gao, Z., Jin, Q., Wang, J., Cao, B. (2020). Clinical features of patients infected with 2019 novel coronavirus in Wuhan, China. Lancet (London, England), 395(10223), 497–506. https://doi.org/10.1016/S0140-6736(20)30183-5

Maguolo, G., Nanni, L. (2020). A Critic Evaluation of Methods for COVID-19 Automatic Detection from XRay Images. arXiv preprint arXiv:2004.12823, 2020.

Ronneberger, O., Fischer, P., Brox, T. U-net: Convolutional networks for biomedical image segmentation. International Conference on Medical image computing and computer-assisted intervention, pages 234–241. Springer, 2015.

World Health Organization (WHO), ed. (May 14th, 2020). Coronavirus disease (COVID-19) Pandemic. https://www.who.int/emergencies/diseases/novel-coronavirus-2019

Wu, F., Zhao, S., Yu, B., Chen, Y. M., Wang, W., Song, Z. G., Hu, Y., Tao, Z. W., Tian, J. H., Pei, Y. Y., Yuan, M. L., Zhang, Y. L., Dai, F. H., Liu, Y., Wang, Q. M., Zheng, J. J., Xu, L., Holmes, E. C., & Zhang, Y. Z. (2020). A new coronavirus associated with human respiratory disease in China. Nature, 579(7798), 265–269. https://doi.org/10.1038/s41586-020-2008-3

